# Assessing psychosis risk using quantitative markers of disorganised speech

**DOI:** 10.1101/2021.01.04.20248717

**Authors:** Sarah E Morgan, Kelly Diederen, Petra E Vértes, Samantha HY Ip, Bo Wang, Bethany Thompson, Arsime Demjaha, Andrea De Micheli, Dominic Oliver, Maria Liakata, Paolo Fusar-Poli, Tom J Spencer, Philip McGuire

**Affiliations:** Department of Computer Science and Technology, University of Cambridge, Cambridge, CB3 0FD, UK; Department of Psychiatry, University of Cambridge, Cambridge, CB2 0SZ, UK; The Alan Turing Institute, London, NW1 2DB, UK; Department of Psychosis Studies, Institute of Psychiatry, Psychology and Neuroscience, King’s College London, London SE5 8AF, UK; Department of Public Health and Primary Care, University of Cambridge, Cambridge, CB1 8RN, UK; Department of Psychiatry, University of Oxford, Oxford, OX3 7JX, United Kingdom; Early Psychosis: Interventions and Clinical-detection (EPIC) lab, Department of Psychosis Studies, Institute of Psychiatry, Psychology and Neuroscience, King’s College London, London SE5 8AF, UK; Department of Brain and Behavioral Sciences, University of Pavia, Pavia, Italy; School of Electronic Engineering and Computer Science, Queen Mary University London, London, E1 4NS, United Kingdom; OASIS service, South London and Maudsley NHS Foundation Trust, London, UK

**Keywords:** Natural language processing, Psychosis, Formal thought disorder, Disorganised speech, Thought and Language Index

## Abstract

Recent work has suggested that disorganised speech might be a powerful predictor of later psychotic illness in clinical high risk subjects. To that end, several automated measures to quantify disorganisation of transcribed speech have been proposed. However, it remains unclear which measures are most predictive of psychosis-onset, how different measures relate to each other and what the best strategies are to elicit disorganised speech from participants. Here, we assessed the ability of twelve automated Natural Language Processing markers to differentiate transcribed speech excerpts from subjects at clinical high risk for psychosis (N=25), first episode psychosis patients (N=16) and healthy control subjects (N=13; N=54 in total). In-line with previous work, several of these measures showed significant differences between groups, including semantic coherence and speech graph connectivity. We also proposed two additional measures of repetition and whether speech was on topic, the latter of which exhibited significant group differences and outperformed the prior, related measure of tangentiality. Most measures examined were only weakly related to each other, suggesting they provide complementary information and that combining different measures could provide additional power to predict the onset of psychotic illness. Finally, we compared the ability of transcribed speech generated using different tasks to differentiate the groups. Speech generated from picture descriptions of the Thematic Apperception Test and a story re-telling task outperformed free speech, suggesting that choice of speech generation method may be an important consideration. Overall, quantitative speech markers represent a promising direction for future diagnostic applications for psychosis risk.

## Introduction

Psychotic disorders affect approximately 1-3% of the population [1] and are among the most disabling of all health conditions. They typically develop at the end of adolescence or in early adulthood, following a clinical high risk (CHR-P) phase, when an individual may or may not progress to frank illness. However, at present it is not possible, on the basis of a clinical assessment, to predict which course a given person at risk is likely to follow [2], [3]. There is thus a clear clinical need to develop methods to quantify an individual’s risk of developing a psychotic disorder, which would allow preventative interventions to be targeted at those who need them most [4].

A core feature of psychotic disorders is Formal Thought Disorder, which is manifest as disorganised or incoherent speech. Recently, several automated approaches have been proposed to quantify speech disorganisation in transcribed speech from patients with psychotic disorders [5]–[11]. For example, Bedi et al [5] introduced an automated measure of speech coherence based on Latent Semantic Analysis (LSA) [12]. Briefly, LSA was used to represent each word as a vector, such that words used in similar contexts (e.g. ‘desk’ and ‘table’) were represented by similar vectors. Semantic coherence was then measured by calculating the average similarity between the words used across a transcribed speech excerpt. Other work [7] used LSA to quantify the tangentiality of an individual’s speech, i.e. how likely it was to diverge off-topic over time. Iter et al [8] found that these measures of semantic coherence and tangentiality could be improved by using new, state-of-the-art word and sentence embedding methods to obtain vectors from words and sentences, instead of LSA. The authors also proposed an automated approach to count the number of times patients used pronouns ambiguously [8], motivated by evidence that patients with schizophrenia may be more likely to use referential pronouns incorrectly [13]. Other authors have used different approaches to quantify disorganised speech, for example Mota et al [10] proposed a graph theoretical approach in which speech was represented as a graph. Speech graph connectivity was significantly reduced in patients with schizophrenia compared to healthy control subjects [10].

These automated approaches allow disorganised speech to be quantified and studied at scale. This is an important improvement on previous qualitative approaches which were subjective and time-consuming, limiting sample sizes. There is also growing evidence that quantitative speech markers might be powerful predictors of later psychotic illness in people at clinical high risk for psychosis. For example, Corcoran et al [6] reported a machine learning classifier which could predict initial psychosis onset with a cross-validated accuracy of approximately 80%, based on decreased semantic coherence (LSA), greater variance in semantic coherence, and reduced usage of possessive pronouns. Similarly, Mota et al [9] obtained approximately 80% accuracy for predicting schizophrenia diagnosis 6 months in advance, based on their speech graph approach [10].

However, most studies only use a limited set of measures to quantify disorganised speech, making it difficult to assess which of these measures are most sensitive and how they are related. It is also unclear which strategies for eliciting speech from participants provide most power to assess thought disorder, for example speech recorded during a task, or free speech recorded from a conversation. This last question is particularly timely given the growing interest in collecting larger samples of speech, with the ultimate aim of predicting disease outcome.

In this study, we first investigated whether twelve Natural Language Processing (NLP) measures could distinguish transcribed speech excerpts from CHR-P subjects, first episode psychosis (FEP) patients and healthy control subjects, using speech excerpts generated by asking participants to describe pictures from the Thematic Apperception Test (TAT; [14]). These pictures typically induce relatively incoherent speech when patients are asked to talk about them in a non-guided way, and have been previously used in conjunction with neuroimaging to identify the neural substrate of abnormal speech in patients with schizophrenia [15], [16]. Ten of the NLP measures employed were taken from the prior literature, and we also developed two additional measures: a measure potentially related to the repetitiveness of speech and a measure of whether a participant’s speech was ‘on-topic’. Second, we investigated whether these NLP measures were correlated with each other, to explore whether they contained overlapping or complementary information. Finally, we assessed whether speech generated using two alternative approaches to the TAT would show similar differences between the three participant groups, to ascertain which strategy for eliciting speech provided most power to assess thought disorder. In particular, we used speech generated by asking participants to re-tell stories from the Discourse Comprehension Test (DCT; [17]) and free speech excerpts.

## Materials and Methods

### Participants

Three groups of participants were recruited as described by Demjaha et al [18]: 25 CHR-P participants, 16 FEP patients and 13 healthy control subjects. CHR-P participants were recruited from the Outreach and Support in South London (OASIS) service [19], which is part of the Pan-London Network for Psychosis-prevention (PNP) [20], and met ultra-high risk criteria assessed with the Comprehensive Assessment of At-Risk Mental States (CAARMS; [21]). FEP patients were recruited from the South London and Maudsley NHS Foundation Trust. Healthy controls with no previous or current history of psychiatric illness and no family history of psychosis were recruited from the same geographical area by local advertisement and by approaching the social contacts of CHR-P individuals after receiving written permission. Healthy controls were matched to the CHR-P individuals and those with FEP for age and gender. Demographics for all three groups are given in **Table 1**.

**Table 1:** Sample characteristics for the three groups: healthy control subjects (CON), clinical high risk subjects (CHR-P) and first episode psychosis patients (FEP). We note that age information was missing for two participants: one CHR-P and one FEP. Results are reported as the mean average and standard deviation where appropriate. WRAT IQ, TLI and education information were missing for one CHR-P subject. TLI; Thought and Language Index. WRAT IQ; Wide Range Achievement Test Intelligence Quotient.

|  | *CON* | *CHR-P* | *FEP* |
| --- | --- | --- | --- |
| *Sample size* | 13 | 25 | 16 |
| *Age (years)* | 26.5 ± 5.2 | 25.1 ± 4.8 | 24.5 ± 3.7 |
| *Sex (M)* | 8 (61.5%) | 15 (60.0%) | 13 (81.3%) |
| *No. on medication* | 0 | 4 | 6 |
| *Years in education* | 18.4 ± 4.2 | 13.0 ± 2.8 | 13.3 ± 1.9 |
| *WRAT IQ* | 115.6 ± 5.2 | 103.3 ± 11.8 | 99.8 ± 15.0 |
| *TLI total* | 0.37 ± 0.51 | 1.8 ± 1.4 | 3.5 ± 2.9 |
| *TLI positive* | 0.37 ± 0.51 | 1.4 ± 1.3 | 2.9 ± 3.0 |
| *TLI negative* | 0 ± 0 | 0.27 ± 0.61 | 0.58 ± 0.86 |

Exclusion criteria for all groups comprised history of a neurological or medical disorder, history of head injury, or alcohol or illicit substance misuse or dependence. All participants were fluent in English and gave written informed consent after receiving a complete description of the study. Ethical approval for the study was obtained from the Institute of Psychiatry Research Ethics Committee.

All CHR-P subjects were followed clinically for an average of 7 years after participating in the study to assess whether they subsequently developed a psychotic disorder. Eight of the 25 CHR-P subjects transitioned to psychosis. Transition to psychosis was defined as the onset of frank psychotic symptoms that did not resolve within a week, corresponding to a severity scale score of 6 on the Disorders of Thought Content subscale, 5 or 6 on the Perceptual Abnormalities subscale and/or 6 on the Disorganized Speech subscales of the CAARMS.

### Procedure

Our primary analyses were performed using transcribed speech generated using the Thematic Apperception Test (TAT; [14]). Participants were presented with eight pictures from the TAT and were asked to talk about each picture for one minute. If the participant stopped talking during the minute they were prompted to continue by the interviewer. Speech samples were recorded and transcribed by a trained assessor who was blinded to participant group status. Parts of speech which were inaudible were noted as [?].

We then repeated the analyses using speech data generated from the same participants with two alternative approaches. In the first, participants were read six stories from the Discourse Comprehension Test (DCT; [17]) and asked to re-tell them to the interviewer, mentioning as many details as possible. Finally, free speech was recorded from an interview with the participants, in which they were asked to speak for 10 minutes about any subject. If the participant stopped talking, they were prompted to continue by the interviewer, using a list of topics the participant was happy to talk about.

We note that data was not available for all participants for all tasks. For the TAT task, no data was available for 1 participant and 1 participant’s recording was excluded due to poor audio quality, leaving N=52. A further 1 participant had 1 picture response (out of 8) missing; this participant was included with only 7 picture descriptions. For the DCT task, 3 participants had no data available, leaving N=51. 6 participants had 1 story response (out of 6) missing and 1 participant had 2 story responses missing; these participants were included with the story responses available. For the free speech, 2 participants had no data available, leaving N=52.

Thought disorder was assessed by applying the Thought and Language Index (TLI; [22]) to speech excerpts from the TAT picture descriptions, again by a trained assessor who was blinded to group status. The positive and negative syndrome scale (PANSS; [23]) was used to measure symptoms in the CHR-P and FEP patient groups. Participants also completed the WRAT IQ test [24] and reported the number of years they spent in education.

### Natural Language Processing measures

#### Basic Measures

For each transcribed speech excerpt, we first calculated the total number of words, *N*_word_, the total number of sentences, *N*_sent_, and the mean number of words per sentence, *N*_word_/*N*_sent_.

#### Semantic coherence

Speech coherence was measured using the same approach as [8]. Briefly, this semantic measure indicates how coherent a text is in terms of the conceptual overlap between adjacent sentences [5]. The text was first split into sentences and pre-processed by removing stop words (defined from the NLTK corpus [25]) and filler words (e.g. ‘ummm’). Each remaining word was then represented as a vector, using word embeddings from the word2vec pre-trained Google News model [26]. From these word embeddings, we then calculated a single vector for each sentence, using Smooth Inverse Frequency (SIF) sentence embedding [27]. We used word2vec and SIF embeddings rather than other available methods because this approach was previously found to give the greatest differences in semantic coherence between patients with schizophrenia and healthy control subjects [8]. Finally, having represented each sentence as a vector, we calculated the cosine similarity between adjacent sentences [5]. The semantic coherence of each response was given by the mean of the cosine similarities between adjacent sentences.

#### Tangentiality

Tangentiality captures the tendency of a subject to drift ‘off-topic’ during discourse. We used the tangentiality measure described by [7], [8], where, for a given response, the cosine similarity was calculated between each sentence in the participant’s response and a prior ‘ground truth’ description of the stimulus used to generate speech (e.g. the TAT picture). As above, we used word2vec and SIF for word and sentence embeddings, respectively. Tangentiality was then computed as the slope of the linear regression of the cosine similarities over time, and can therefore range from −1 to 1. A more negative slope means the response became less closely related to the stimulus as time progresses.

To calculate tangentiality for the TAT task, we used prior descriptions of each of the 8 pictures from [28], given for reference in **Supplementary Section S1**. For the DCT story retell task we used the original stories to calculate the ground truth vectors with which the participants’ responses were compared. Note that we did not obtain tangentiality scores for the free speech conversations, due to the absence of a ground truth for the conversations.

#### On-topic score

We propose an ‘on-topic’ score, which is closely related to tangentiality. Here, instead of calculating the slope of the cosine similarities over time, we calculated the mean of the cosine similarities between each sentence and the ground truth stimulus description (again ranging from −1 to 1). This measure captures how ‘on-topic’ the participant’s response to the stimulus was on average across the whole response, rather than whether it became less closely related to the stimulus over time. As above, we used the TAT picture descriptions from [28] and the original DCT stories as the ground truth stimulus descriptions, and we did not obtain on-topic scores for free speech.

#### Repetition

Prior work has suggested that speech from patients with schizophrenia may be more repetitive than speech from healthy control subjects [22]. As a first step towards measuring the repetitiveness of speech in a quantitative way, we calculated the cosine similarity between all possible pairs of sentences, and defined a candidate repetition score as the maximum cosine similarity between any two sentences (ranging from −1 to 1). A maximum similarity score of 1 means that (at least) two of the sentences in the response were represented by identical vectors, suggesting the same content was repeated.

#### Number of ambiguous pronouns

Given evidence that patients with schizophrenia may not use referential pronouns correctly [13], [8] proposed to count the number of ambiguous pronouns as a syntactic measure of speech incoherence. Here, ambiguous pronouns are pronouns which were either 1) never resolved (e.g. ‘‘I think that’s *their* dog’’, where ‘‘they’’ are never named) or 2) resolved only after the use of a proper noun (e.g. ‘‘I told *him* to go away, *my friend*, I didn’t want to see him’’) [8]. Following [8], we first identified all the pronouns in a participant’s response and the subject they referred to, using a pre-trained co-reference resolution model [29]. We then counted the number of times the first term used to refer to a subject was a third-person pronoun (he, she etc).

#### Speech graphs

Speech graphs were proposed by [11] and provide an alternative approach to quantify speech incoherence. Briefly, each unique word in a participant’s response is represented by a node, and directed edges link the words in the order in which they were spoken. We note that prior work has already applied speech graph analysis to our TAT speech excerpts [30], and found significant group differences in speech graph connectivity measures. Here, we compared speech graph connectivity to the other NLP measures described above. We also applied the speech graph approach to speech excerpts obtained using the DCT task, and free speech.

As described by [30], we used the SpeechGraphs software [10] to calculate two measures of graph connectivity: the total number of nodes in the largest connected component (LCC) and the largest strongly connected component (LSC) [9], [10]. The LCC is the largest sub-graph in which all nodes are linked by at least one path. The LSC is the largest sub-graph in which all nodes are linked by a path which can be traversed in either direction (i.e. there is a path from node *i* to node *j*, and also from node *j* to node *i*). To control for the number of words spoken, LCC and LSC were calculated for windows of 30 words, which overlapped by 15 words, then averaged across all windows [30]. We also calculated values of LCC and LSC normalised to the equivalent measures from randomised speech graphs (obtained by randomly shuffling the words within each window), to determine how close to randomness the connectedness measures were; denoted LCCr and LSCr [30].

### Statistical Analyses

The metrics described above were calculated for each speech excerpt, for each subject. Where there was more than one speech excerpt available per subject (e.g. for the TAT, each subject was asked to describe 8 pictures, giving 8 excerpts), we calculated the mean of the scores across the excerpts, to obtain a single value per subject.

We used the Shapiro-Wilk test to assess the Normality of the NLP measures, see **Supplementary Section S2**. Some measures were not Normally distributed, and we used the Mann-Whitney U-test to calculate the statistical significance of group differences. The relationships between different NLP measures, and between the NLP measures and the TLI, symptom scores and cognitive measures, were calculated by performing linear regressions, controlling for group membership as a co-variate.

Finally, we counted the number of inaudible pieces of speech in each excerpt, normalised to the total number of words in the excerpt. We assessed whether there were significant differences in the number of inaudible pieces of speech per word between groups or between methods used to generate speech (TAT/DCT/free speech) using the Mann-Whitney U-test. For those methods where there were differences, as an additional sensitivity analysis we tested whether group differences in the NLP metrics remained significant when controlling for the number of inaudible pieces of speech per word. To that end we used a Generalized Additive Model for Location, Scale and Shape (GAMLSS) with a gamma distribution [31].

## Results

### Speech profiles

We first calculated all twelve NLP measures outlined in the Methods section, for the TAT excerpts from all subjects. The average values for all measures per group are shown as average ‘speech profiles’ (spider plots) in **Figure 1A**. For illustrative purposes, we also show speech profiles for two individual participants’ descriptions of the picture in **Figure 1B**. The CHR-P subject’s speech profile shown in **Figure 1C** exhibits a somewhat higher number of sentences and reduced number of words compared to the average control subject. The semantic coherence score, on-topic score and speech graph measures are also lower than those for the average control. The control subject’s speech profile shown in **Figure 1D** follows the average control subject’s speech profile more closely, but does exhibit a higher maximum similarity (repetition) score.

**Figure 1:**
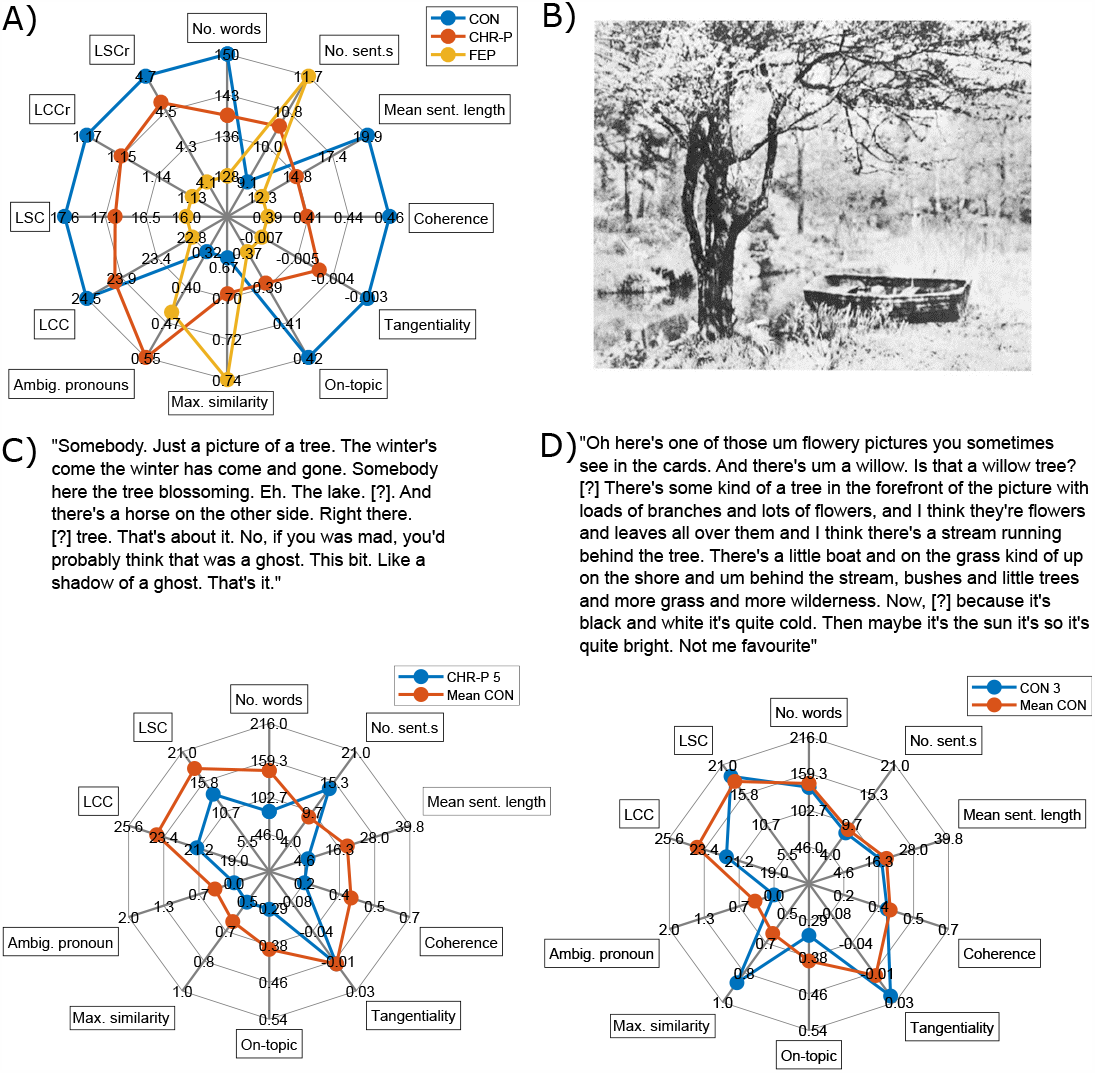
Speech profiles. A) Average speech profiles for the control subjects, CHR-P subjects and FEP patients. B) One of the images used from the TAT. Example descriptions of this picture are shown in C) and D) for a particular CHR-P and control subject, respectively. The response in part C) diverges somewhat from the average control response, with more, shorter sentences, and lower coherence, on-topic score and LCC, for example. The response in part D) follows the average control response quite closely, but has a somewhat higher maximum similarity between sentences. We note that the healthy control subject whose speech profile is given in part D) was excluded from our calculation of the average control response, to avoid inflating the similarity between their speech profile and the average control profile. Spider plots were generated using code from [33].

### Group differences in NLP measures, for the TAT

Group differences for all NLP measures obtained from the TAT speech excerpts are given in **Table 2**, with corresponding box-plots shown in **Figure 2**. Comparing FEP patients to control subjects, both number of words and mean sentence length were significantly lower for FEP patients, whilst the number of sentences was significantly higher. We also observed lower semantic coherence for FEP patients, in-line with results from [8]. The measure of tangentiality from [8] did not show any significant group differences, however our on-topic score significantly decreased in FEP patients, showing a larger group difference than any other measure. This suggests that FEP patients’ responses did not diverge from the prior picture description over time, but were instead less closely related to the prior picture description on average across all time points. There were no significant differences in the ambiguous pronoun count between the FEP patients and healthy control subjects, in contrast to results obtained by [8]. The maximum similarity (repetition) measure showed no statistically significant group differences. As previously reported by [30], there were significant reductions in speech graph connectivity in FEP patients, in-line with [9], [10].

**Table 2:**
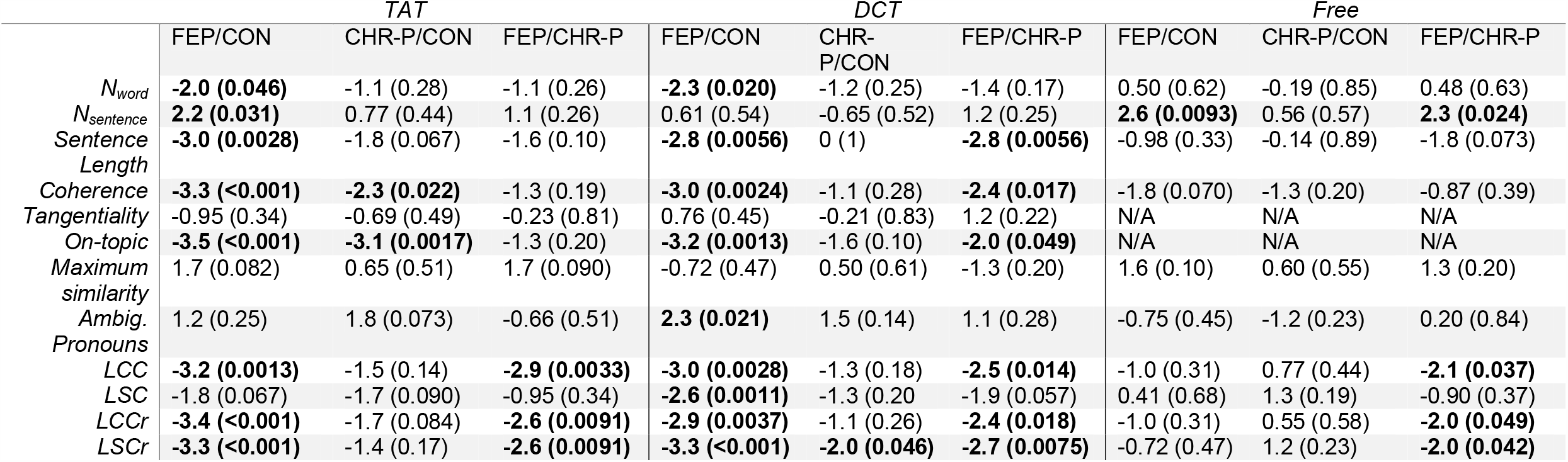
Statistical group differences in NLP measures. Z-values are given from Mann-Whitney U-tests, with the corresponding P-values in brackets. LCC, LSC, LCCr and LSCr results for the TAT have already been reported by [30].

|  | TAT |  |  | DCT |  |  | Free |  |  |
| --- | --- | --- | --- | --- | --- | --- | --- | --- | --- |
|  | FEP/CON | CHR-P/CON | FEP/CHR-P | FEP/CON | CHR-P/CON | FEP/CHR-P | FEP/CON | CHR-P/CON | FEP/CHR-P |
| *N<sub>word</sub>* | **-2.0 (0.046)** | -1.1 (0.28) | -1.1 (0.26) | **-2.3 (0.020)** | -1.2 (0.25) | -1.4 (0.17) | 0.50 (0.62) | -0.19 (0.85) | 0.48 (0.63) |
| *N<sub>sentence</sub>* | **2.2 (0.031)** | 0.77 (0.44) | 1.1 (0.26) | 0.61 (0.54) | -0.65 (0.52) | 1.2 (0.25) | **2.6 (0.0093)** | 0.56 (0.57) | **2.3 (0.024)** |
| *Sentence Length* | **-3.0 (0.0028)** | -1.8 (0.067) | -1.6 (0.10) | **-2.8 (0.0056)** | 0 (1) | **-2.8 (0.0056)** | -0.98 (0.33) | -0.14 (0.89) | -1.8 (0.073) |
| *Coherence* | **-3.3 (<0.001)** | **-2.3 (0.022)** | -1.3 (0.19) | **-3.0 (0.0024)** | -1.1 (0.28) | **-2.4 (0.017)** | -1.8 (0.070) | -1.3 (0.20) | -0.87 (0.39) |
| *Tangentiality* | -0.95 (0.34) | -0.69 (0.49) | -0.23 (0.81) | 0.76 (0.45) | -0.21 (0.83) | 1.2 (0.22) | N/A | N/A | N/A |
| *On-topic* | **-3.5 (<0.001)** | **-3.1 (0.0017)** | -1.3 (0.20) | **-3.2 (0.0013)** | -1.6 (0.10) | **-2.0 (0.049)** | N/A | N/A | N/A |
| *Maximum similarity* | 1.7 (0.082) | 0.65 (0.51) | 1.7 (0.090) | -0.72 (0.47) | 0.50 (0.61) | -1.3 (0.20) | 1.6 (0.10) | 0.60 (0.55) | 1.3 (0.20) |
| *Ambig. Pronouns* | 1.2 (0.25) | 1.8 (0.073) | -0.66 (0.51) | **2.3 (0.021)** | 1.5 (0.14) | 1.1 (0.28) | -0.75 (0.45) | -1.2 (0.23) | 0.20 (0.84) |
| *LCC* | **-3.2 (0.0013)** | -1.5 (0.14) | **-2.9 (0.0033)** | **-3.0 (0.0028)** | -1.3 (0.18) | **-2.5 (0.014)** | -1.0 (0.31) | 0.77 (0.44) | **-2.1 (0.037)** |
| *LSC* | -1.8 (0.067) | -1.7 (0.090) | -0.95 (0.34) | **-2.6 (0.0011)** | -1.3 (0.20) | -1.9 (0.057) | 0.41 (0.68) | 1.3 (0.19) | -0.90 (0.37) |
| *LCCr* | **-3.4 (<0.001)** | -1.7 (0.084) | **-2.6 (0.0091)** | **-2.9 (0.0037)** | -1.1 (0.26) | **-2.4 (0.018)** | -1.0 (0.31) | 0.55 (0.58) | **-2.0 (0.049)** |
| *LSCr* | **-3.3 (<0.001)** | -1.4 (0.17) | **-2.6 (0.0091)** | **-3.3 (<0.001)** | **-2.0 (0.046)** | **-2.7 (0.0075)** | -0.72 (0.47) | 1.2 (0.23) | **-2.0 (0.042)** |

**Figure 2:**
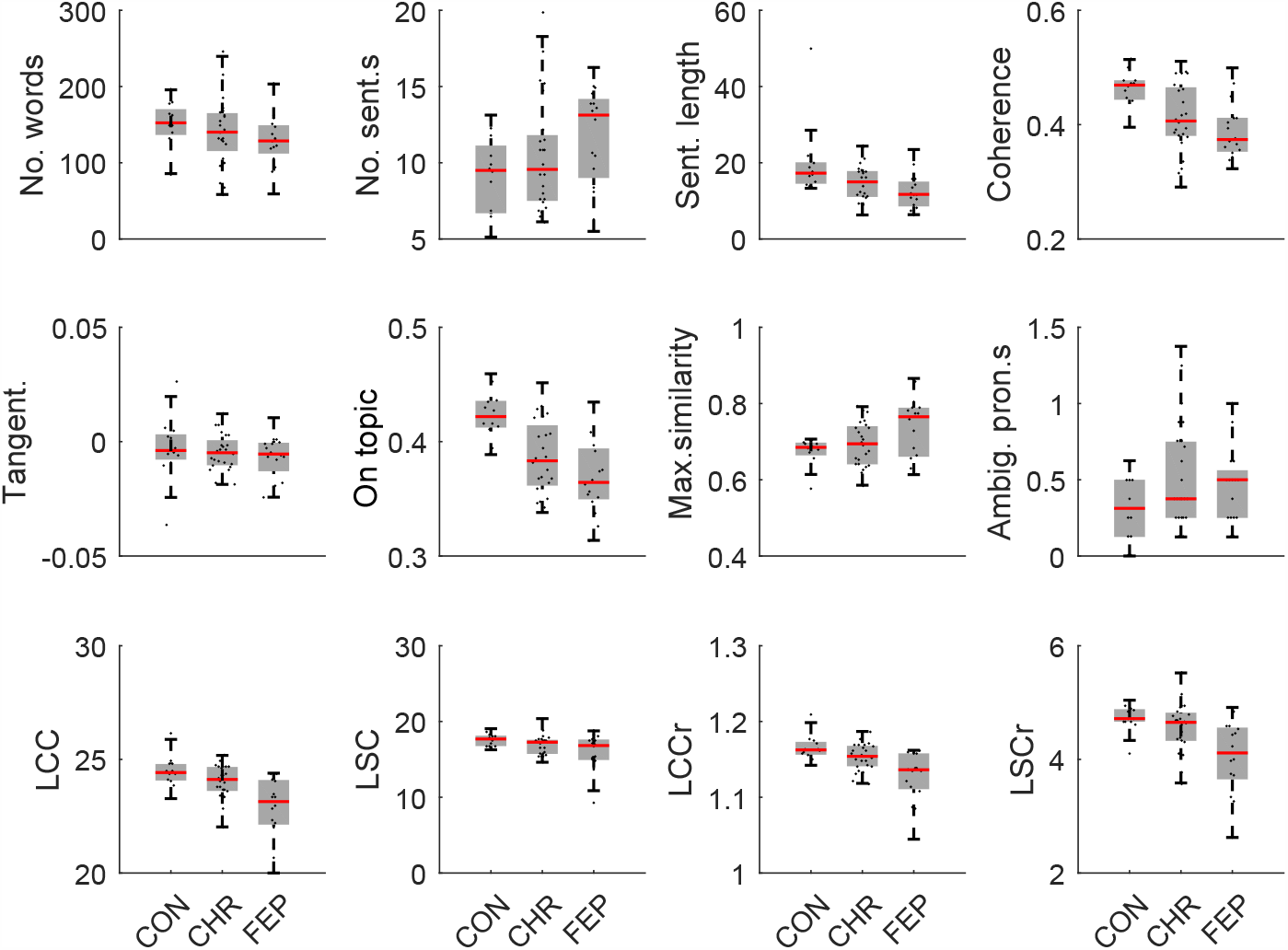
Box-plots showing group differences in all twelve NLP measures, for speech generated using the TAT.

In the CHR-P group, on-topic score and semantic coherence were reduced compared to the healthy control subjects. These measures showed no significant differences between CHR-P subjects and FEP patients. In contrast, LCC, LCCr and LSC increased in CHR-P subjects with respect to FEPs, but showed no significant differences between CHR-P subjects and the control group.

As shown in **Table 1**, 4 of the CHR-P subjects and 6 of the FEP patients were taking antipsychotic medication. Excluding subjects who were taking medication did not qualitatively change the group differences in the NLP measures; see **Supplementary Section S4**.

We did not observe significant differences in any of the twelve NLP measures between the 8 CHR-P subjects who subsequently transitioned to psychosis and the 16 CHR-P subjects who did not transition; see **Supplementary Section S5**.

### Inaudible pieces of speech

For the TAT speech excerpts, there were no significant differences in the number of inaudible pieces of speech per word between the FEP patients and the control subjects (*Z* = 1.3, *P* = 0.20), or between the FEP patients and the CHR-P subjects (*Z* = −0.95, *P* = 0.34). However, there was a significant difference in the number of inaudible pieces of speech per word between the CHR-P subjects and the healthy control subjects (*Z* = 2.5, *P* = 0.014). All previously identified group differences in NLP metrics remained significant when controlling for the number of inaudible pieces of speech per word, apart from the decrease in total number of words observed in the FEP patients compared to the healthy controls (*Z* = −1.8, *P* = 0.084); see **Supplementary Section S6**.

### Relationships between NLP measures

We next explored whether the NLP measures were significantly associated with each other, by fitting a linear regression model to each pair of NLP measures, controlling for group as a co-variate. **Figure 3 A**) shows the relationships between the NLP measures, with those that were significant with *P* < 0.01 plotted in the network in **Figure 3 B**).

**Figure 3:**
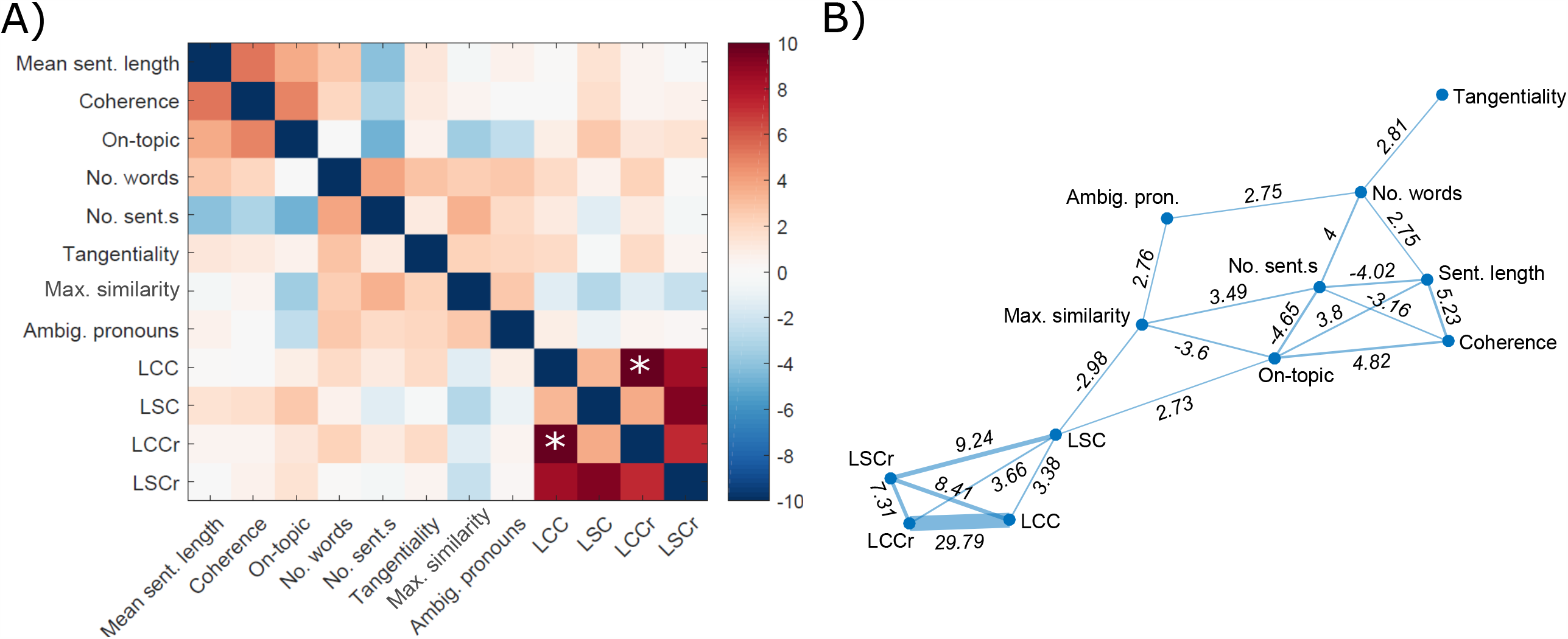
Relationships between NLP measures. A) Heat mapping showing the relationships (T-statistics) between different NLP measures, calculated using linear regression, controlling for group membership. Colormap from [34]. B) Network showing the NLP measures which are significantly associated with each other, with *P*<0.01. Corresponding T-statistics are shown on the network edges between measures. *The colorbar was truncated at *T*=10 for visualisation purposes; *T*=29.79 for the relationship between LCC and LCCr.

The four speech graph measures (LCC, LCCr, LSC and LSCr) were strongly associated with each other, as expected. There was also a significant negative association between LSC and maximum similarity (repetition), and a significant positive association between LSC and on-topic score. Interestingly, there was no significant association between any of the speech graph measures and semantic coherence. Semantic coherence was significantly negatively associated with number of sentences and significantly positively associated with sentence length and on-topic score.

### Relationships between NLP measures and the TLI, symptoms and cognitive measures

All subjects had data available for the TLI, IQ and number of years in education, whilst 15 CHR-P subjects, 8 FEP patients and no control subjects had PANSS data available. **Supplementary Table S2** shows the associations between the NLP measures and the TLI (TLI total, TLI positive and TLI negative), PANSS symptoms (PANSS positive, PANSS negative and PANSS general), IQ and number of years in education. After FDR correction for multiple comparisons (12 x 8 = 96 comparisons across all NLP and TLI, symptom and cognitive measures), we observed significant associations between TLI negative and: the number of words (*T*=-4.7, *P*_*FDR*_=0.0010), LCC (*T*=-4.1, *P*_*FDR*_=0.0038), LCCr (*T*=-5.4, *P*_*FDR*_<0.001), LSC (*T*=-4.4, *P*_*FDR*_=0.0023) and LSCr (*T*=-3.6, *P*_*FDR*_=0.014).

### DCT task and free speech

Finally, we re-calculated the group differences for each of the NLP measures using speech generated from either the DCT story retelling task or free speech. Results are shown in **Table 2**. With the DCT task, we observed a significant decrease in semantic coherence and on-topic score in FEP patients with respect to healthy controls, as well as in the number of words, mean sentence length, LCC, LCCr and LSCr, replicating the equivalent results for the TAT task. All of these measures apart from number of words and LCCr also showed significant reductions in FEP patients with respect to CHR-P subjects, but there were no significant differences between CHR-P subjects and healthy control subjects apart from for LSCr (unlike the TAT task where semantic coherence and on-topic score showed significant differences between CHR-P and control subjects, but not between CHR-P and FEP patients). With the DCT task we also observed a significant increase in the number of ambiguous pronouns in FEP patients with respect to control subjects, but there was no difference in ambiguous pronoun count between CHR-P subjects and either FEP patients or healthy controls.

With free speech, we observed a significant increase in the number of sentences spoken by FEP patients with respect to both CHR-P subjects and healthy controls. However, none of the other measures showed significant differences between FEP patients and healthy control subjects, including semantic coherence, on-topic score and maximum similarity. We note that the maximum similarity measure gave the highest possible score of 1 for several of the free speech excerpts, unlike for the TAT and DCT. This was due to the greater length of the free speech excerpts compared to the TAT and DCT excerpts, and suggests the measure may need adapting for use with longer excerpts. Interestingly, we did observe a significant decrease in LCC, LCCr and LSCr in FEP patients with respect to CHR-P subjects, despite there being no significant difference between these measures for FEP patients and healthy controls.

There were no group differences in number of inaudible pieces of speech per word for the free speech excerpts, although there was a significant increase in number of inaudible pieces of speech per word for the FEP patients compared to control subjects for the DCT speech excerpts (*Z*=2.0, *P*=0.047). All previously identified group differences in NLP metrics observed from the DCT excerpts remained significant when controlling for the number of inaudible pieces of speech per word with the GAMLSS model, apart from the decrease in total number of words observed in the FEP patients compared to the healthy controls which was no longer significant (*Z* = −0.28, *P* = 0.78), and the difference in ambiguous pronoun count between FEP patients and healthy controls, which we were not able to test with the GAMLSS model; see **Supplementary Section S6** for details.

Whilst there was no significant difference in number of inaudible pieces of speech per word between the TAT and DCT speech excerpts, we did observe a significant reduction in number of inaudible pieces of speech per word in the free speech excerpts compared to both the TAT (*Z*=-3.4, *P*<0.001) and the DCT excerpts (*Z*=-4.0, *P*<0.001), see **Supplementary Section S6**.

## Discussion

Our primary analysis of the TAT picture speech excerpts showed that several NLP measures did indeed discriminate between groups. Notably, from the prior literature both semantic coherence [8] and speech graph connectivity [10], [11] were significantly reduced in FEP patients compared to control subjects. Semantic coherence and speech graph connectivity also distinguished CHR-P subjects from control subjects and FEP patients, respectively. Ambiguous pronoun count did not show significant group differences, but may be worth revisiting with more accurate co-reference resolution models as they become available. We additionally proposed two measures to capture how ‘on-topic’ and repetitive speech was. Whilst there were no significant group differences in repetition (measured as maximum similarity between pairs of sentences), the on-topic score exhibited significant group differences between control subjects and both CHR-P subjects and FEP patients. This was in contrast to the related measure of tangentiality [7], [8], which did not exhibit any significant group differences. We therefore believe the on-topic measure merits inclusion in future research studies.

Second, we investigated the relationships between different NLP measures. There were some significant correlations, for example we observed a negative correlation between LSC speech graph connectivity and the maximum similarity measure, which makes sense given that repetitive speech with fewer unique words will lead to fewer nodes being included in a speech graph and hence reduced connectivity. Our ‘on-topic’ measure was also significantly positively correlated with semantic coherence and the LSC speech graph connectivity. Nonetheless, most inter-measure relationships were weak, for example there was no significant correlation between speech graph connectivity and semantic coherence.

These results suggest that different NLP measures may provide complementary information. It is predictable that different speech measures may capture distinct aspects of psychosis, e.g. different symptoms. It also seems likely that combining different measures in machine learning algorithms might give additional power to predict future disease trajectories for CHR-P subjects, compared to using a single measure. Future studies should examine multiple NLP measures concurrently in larger samples, to test these hypotheses. The low computational cost of calculating the automated NLP measures described in this paper (at most seconds per participant) makes extracting multiple measures computationally straightforward.

Finally, we explored the impact of using different approaches to generate speech. Speech generated using the DCT story task replicated many of the NLP group differences observed with the TAT pictures. Free speech exhibited fewer, weaker NLP group differences compared to speech generated using the TAT pictures or the DCT story task, suggesting that this approach may be less sensitive for assessing thought disorder, although there were more inaudible pieces of speech per word in the free speech excerpts and we cannot rule out the possibility this contributed to the weaker NLP group differences (see Limitations). A task-dependency is in-line with previous work, which found speech in which participants described their dreams was more predictive of psychosis than speech in which participants described their waking activities [10]. We were also unable to generate all NLP measures from free speech excerpts, for example due to a lack of ‘ground-truth’ stimulus description from which to calculate on-topic scores. These observations suggest that the task(s) used to generate speech in future studies should be considered carefully.

## Limitations

Ultimately, further external work is required before speech measures are ready to be “rolled out” to clinical applications.

A key limitation of this study was the sample size, which was in-line with prior work, but still small considering the known heterogeneity of CHR-P subjects [32]. In addition, the particularly small number of CHR-P subjects who transitioned to psychosis (N=8) may mean the absence of significant differences in NLP measures between CHR-P subjects who did and did not transition was due to a lack of statistical power. It will therefore be important for future work to externally validate our results to test their generalisability, and to examine larger cohorts of CHR-P subjects who did and did not transition to psychosis. We will make our code openly available on publication, to enable other researchers to implement the same approaches.

The modest sample size also meant that we focussed on group-level, statistical analyses. However, to be clinically useful, future work will need to use NLP measures to predict disease outcomes for individual patients, for example by applying more “data hungry” machine learning approaches to larger samples. We believe that the results of this study provide an important step towards large studies at the individual level, by highlighting which methods may be best suited to generating speech in future studies and the potential power of combining multiple NLP measures when analysing data. When turning to how disorganised speech manifests at the individual level, across measures, visualisation tools like the speech profiles proposed here could prove particularly valuable.

Finally, group comparison studies are vulnerable to differences in confounding factors between groups and here there were group differences in antipsychotic medication, IQ and number of years of education (**Table 1**). Excluding subjects who had been prescribed antipsychotic medication did not qualitatively change our main results (**SI Section S4**). We also did not observe significant associations between any of the NLP measures and IQ or number of years in education (**SI Section S3**). Nonetheless, we cannot completely rule out the possibility that these or other, unobserved confounding factors might contribute to differences in NLP measures between groups. There were also significantly more inaudible pieces of speech per word in the free speech excerpts compared to the TAT and DCT excerpts, which may be related to the weaker group differences in NLP metrics observed in the free speech excerpts.

## Conclusions

Overall, automated approaches to assessing disorganised speech show substantial promise for diagnostic applications. Quantifying incoherent speech may also give fresh insights into how this core symptom of psychotic disorders manifests.

## Data Availability

Data is not available due to ethical reasons (participants did not agree for their data to be shared publicly). Code will be made available on publication.

## Acknowledgements

We thank the services users and volunteers who took part in this study, and the members of the Outreach and Support in South London (OASIS) team who were involved in the recruitment, management and clinical follow-up of the participants reported in this manuscript. We are also grateful to the experts by experience from the Cambridge and Peterborough Foundation Trust Service User and Carers Research Group, who gave constructive feedback on the manuscript. SEM was funded by a Fellowship from The Alan Turing Institute, London, and a Henslow Fellowship at Lucy Cavendish College, University of Cambridge, funded by the Cambridge Philosophical Society. PEV is supported by a fellowship from MQ: Transforming Mental Health (MQF17_24). This work was supported by The Alan Turing Institute under the EPSRC grant EP/N510129/1, the UK Medical Research Council (MRC) and the National Institute for Health Research (NIHR) Mental Health Biomedical Research Centre at South London and Maudsley NHS Foundation Trust and King’s College London. The views expressed are those of the author(s) and not necessarily those of the NHS, the NIHR, MRC or the Department of Health. The funder had no influence on the design of the study or interpretation of the results.

## Conflicts of interest

The authors declare that the research was conducted in the absence of any commercial or financial relationships that could be construed as a potential conflict of interest.

## Notes

### Competing Interest Statement

The authors have declared no competing interest.

### Author Declarations

Ethical approval for the study was obtained from the Institute of Psychiatry Research Ethics Committee at King's College London.

## References

[1] J. Perälä et al., “Lifetime Prevalence of Psychotic and Bipolar I Disorders in a General Population,” Arch. Gen. Psychiatry, vol. 64, no. 1, p. 19, Jan. 2007, doi:10.1001/archpsyc.64.1.19.

[2] P. Fusar-Poli, S. A. Sullivan, J. L. Shah, and P. J. Uhlhaas, “Improving the detection of individuals at clinical risk for psychosis in the community, primary and secondary care: An integrated evidence-based approach,” Front. Psychiatry, vol. 10, no. OCT, 2019, doi:10.3389/fpsyt.2019.00774.

[3] D. Prata, A. Mechelli, and S. Kapur, “Clinically meaningful biomarkers for psychosis: A systematic and quantitative review,” Neuroscience and Biobehavioral Reviews. 2014, doi:10.1016/j.neubiorev.2014.05.010.

[4] P. Fusar-Poli et al., “Prevention of Psychosis: Advances in Detection, Prognosis, and Intervention,” JAMA Psychiatry, vol. 77, no. 7. American Medical Association, pp. 755–765, Jul. 01, 2020, doi:10.1001/jamapsychiatry.2019.4779.

[5] G. Bedi et al., “Automated analysis of free speech predicts psychosis onset in high-risk youths,” npj Schizophr., 2015, doi:10.1038/npjschz.2015.30.

[6] C. M. Corcoran et al., “Prediction of psychosis across protocols and risk cohorts using automated language analysis,” World Psychiatry, 2018, doi:10.1002/wps.20491.

[7] B. Elvevåg, P. W. Foltz, D. R. Weinberger, and T. E. Goldberg, “Quantifying incoherence in speech: An automated methodology and novel application to schizophrenia,” Schizophr. Res., vol. 93, no. 1–3, pp. 304–316, Jul. 2007, doi:10.1016/J.SCHRES.2007.03.001.

[8] D. Iter, J. H. Yoon, and D. Jurafsky, “Automatic Detection of Incoherent Speech for Diagnosing Schizophrenia,” 2018. Accessed: Jun. 24, 2019. [Online]. Available: https://www.aclweb.org/anthology/W18-0615.

[9] N. B. Mota, M. Copelli, and S. Ribeiro, “Thought disorder measured as random speech structure classifies negative symptoms and schizophrenia diagnosis 6 months in advance,” npj Schizophr., vol. 3, no. 1, p. 18, Dec. 2017, doi:10.1038/s41537-017-0019-3.

[10] N. B. Mota, R. Furtado, P. P. C. Maia, M. Copelli, and S. Ribeiro, “Graph analysis of dream reports is especially informative about psychosis,” Sci. Rep., vol. 4, no. 1, pp. 1–7, Jan. 2014, doi:10.1038/srep03691.

[11] N. B. Mota et al., “Speech Graphs Provide a Quantitative Measure of Thought Disorder in Psychosis,” PLoS One, vol. 7, no. 4, p. e34928, Apr. 2012, doi:10.1371/journal.pone.0034928.

[12] T. K. Landauer, P. W. Foltz, and D. Laham, “An introduction to latent semantic analysis,” Discourse Process., vol. 25, no. 2–3, pp. 259–284, Jan. 1998, doi:10.1080/01638539809545028.

[13] W. Hinzen, “The linguistics of schizophrenia: thought disturbance as language pathology across positive symptoms,” Front. Psychol., vol. 6, 2015, doi:10.3389/fpsyg.2015.00971.

[14] H. Murray, Thematic Apperception Test. Harvard University Press, 1943.

[15] P. K. McGuire, D. J. Quested, S. A. Spence, R. M. Murray, C. D. Frith, and P. F. Liddle, “Pathophysiology of ‘positive’ thought disorder in schizophrenia.,” Br. J. Psychiatry, vol. 173, pp. 231–5, Sep. 1998, Accessed: Jun. 28, 2019. [Online]. Available: http://www.ncbi.nlm.nih.gov/pubmed/9926099.

[16] T. T. Kircher, P. F. Liddle, M. J. Brammer, S. C. Williams, R. M. Murray, and P. K. McGuire, “Neural correlates of formal thought disorder in schizophrenia: preliminary findings from a functional magnetic resonance imaging study.,” Arch. Gen. Psychiatry, vol. 58, no. 8, pp. 769–74, Aug. 2001, Accessed: Jun. 28, 2019. [Online]. Available: http://www.ncbi.nlm.nih.gov/pubmed/11483143.

[17] R. Brookshire and L. Nicholas, “The discourse comprehension test.” Tucson, AZ: Communication Skill Builders/The Psychological Corporation, 1993.

[18] A. Demjaha et al., “Formal thought disorder in people at ultra-high risk of psychosis.,” BJPsych open, vol. 3, no. 4, pp. 165–170, Jul. 2017, doi:10.1192/bjpo.bp.116.004408.

[19] P. Fusar-Poli, M. Byrne, S. Badger, L. R. Valmaggia, and P. K. McGuire, “Outreach and support in South London (OASIS), 2001-2011: Ten years of early diagnosis and treatment for young individuals at high clinical risk for psychosis,” Eur. Psychiatry, vol. 28, no. 5, pp. 315–326, Jun. 2013, doi:10.1016/j.eurpsy.2012.08.002.

[20] P. Fusar-Poli et al., “Pan-London Network for Psychosis-Prevention (PNP),” Front. Psychiatry, vol. 10, Oct. 2019, doi:10.3389/fpsyt.2019.00707.

[21] A. R. Yung et al., “Mapping the Onset of Psychosis: The Comprehensive Assessment of At-Risk Mental States,” Aust. New Zeal. J. Psychiatry, vol. 39, no. 11–12, pp. 964–971, Nov. 2005, doi:10.1080/j.1440-1614.2005.01714.x.

[22] P. F. Liddle et al., “Thought and Language Index: an instrument for assessing thought and language in schizophrenia.,” Br. J. Psychiatry, vol. 181, pp. 326–30, Oct. 2002, Accessed: Jun. 24, 2019. [Online]. Available: http://www.ncbi.nlm.nih.gov/pubmed/12356660.

[23] S. R. Kay, A. Flszbeln, and L. A. Qpjer, “The Positive and Negative Syndrome Scale (PANSS) for Schizophrenia,” 1967. Accessed: Jul. 01, 2020. [Online]. Available: https://academic.oup.com/schizophreniabulletin/article-abstract/13/2/261/1919795.

[24] G. Wilkinson and G. Robertson, WRAT 4□: wide range achievement test professional manual, 4th ed. Lutz FL: Psychological Assessment Resources Inc., 2006.

[25] S. Bird, E. Klein, and E. Loper, Natural Language Processing with Python. O’Reilly Media, Inc., 2009.

[26] T. Mikolov, K. Chen, G. Corrado, and J. Dean, “Efficient estimation of word representations in vector space,” Jan. 2013, Accessed: Jul. 01, 2020. [Online]. Available: http://ronan.collobert.com/senna/.

[27] S. Arora, Y. Liang, and T. Ma, “A SIMPLE BUT TOUGH-TO-BEAT BASELINE FOR SEN-TENCE EMBEDDINGS.” Accessed: Jul. 01, 2020. [Online]. Available: https://github.com/PrincetonML/SIF.

[28] P. Shrestha, “Detailed Procedure of Thematic Apperception test - Psychestudy.” https://www.psychestudy.com/general/personality/detailed-procedure-thematic-procedure-test (Accessed Jul. 01, 2020).

[29] K. Lee, L. He, M. Lewis, and L. Zettlemoyer, “End-to-end Neural Coreference Resolution,” EMNLP 2017 - Conf. Empir. Methods Nat. Lang. Process. Proc., pp. 188–197, Jul. 2017, Accessed: Jul. 01, 2020. [Online]. Available: http://arxiv.org/abs/1707.07045.

[30] T. J. Spencer et al., “Lower speech connectedness linked to incidence of psychosis in people at clinical high risk,” Schizophr. Res., Sep. 2020, doi:10.1016/j.schres.2020.09.002.

[31] R. A. Rigby, D. M. Stasinopoulos, and P. W. Lane, “Generalized additive models for location, scale and shape,” J. R. Stat. Soc. Ser. C Appl. Stat., vol. 54, no. 3, pp. 507–554, Jun. 2005, doi:10.1111/j.1467-9876.2005.00510.x.

[32] P. Fusar-Poli et al., “Heterogeneity of psychosis risk within individuals at clinical high risk: A meta-analytical stratification,” JAMA Psychiatry, vol. 73, no. 2, pp. 113–120, Feb. 2016, doi:10.1001/jamapsychiatry.2015.2324.

[33] “NewGuy012/spider_plot: Create a spider or radar plot with individual axes.” https://github.com/NewGuy012/spider_plot (Accessed Jul. 10, 2020).

[34] “cbrewer□: colorbrewer schemes for Matlab - File Exchange - MATLAB Central.” https://www.mathworks.com/matlabcentral/fileexchange/34087-cbrewer-colorbrewer-schemes-for-matlab (Accessed Jul. 10, 2020).

