## Supplementary Information for "Assessing psychosis risk using quantitative markers of disorganised speech"

December 22, 2020

#### Contents

1 TAT Picture Descriptions 2

2 Normality 2

3 Relationships between NLP measures and the TLI, PANSS scores and cognitive measures 2

4 Medication 4

5 Transition to psychosis 4

6 Inaudible pieces of speech 4

*TJS and PM share joint last authorship

### 1 TAT Picture Descriptions

TAT picture descriptions were taken from [1], and are copied below for reference.

**Picture 6GF:** An older man with a pipe in his mouth is talking to a younger woman sitting on a couch, who is looking back at him.

**Picture 7GF:** A young girl is sitting on a couch with a doll in her hands, and an older woman sitting behind her is reading to her from a book.

**Picture 16B:** Three hands are grabbing a man dressed in a long coat.

**Picture 13B:** A boy sitting in the doorway of a log cabin.

**Picture 9BM:** Four men lying in a field against one another.

**Picture 15:** A man is standing with his hands clasped together. There are tombstones everywhere.

**Picture 12BG:** There is a tree and a rowboat next to it in a country setting with no presence of a human being.

**Picture 20:** The card shows a man leaning against a lamppost at night in a hazy atmosphere.

### 2 Normality

We performed Shapiro-Wilk tests to test the Normality of the NLP measures, for the TAT speech excerpts. Results are shown in Table S1.

| NLP measure | Controls | CHR-P | FEP |
|---|---|---|---|
| No. words | 0.74 | 0.67 | 0.95 |
| No. sent.s | 0.61 | **0.021** | 0.17 |
| Mean sent. length | **<0.001** | 0.32 | 0.24 |
| Coherence | 0.74 | 0.36 | 0.21 |
| Tangentiality | 0.12 | 0.90 | 0.21 |
| On-topic | 0.91 | 0.33 | 0.69 |
| Max. similarity | **0.0088** | 0.47 | 0.22 |
| Ambig. Pronouns | 0.21 | **0.021** | 0.15 |
| LCC | 0.40 | 0.23 | 0.16 |
| LSC | 0.73 | 0.16 | **0.030** |
| LCCr | 0.099 | 0.81 | **0.039** |
| LSCr | **0.033** | 0.78 | 0.45 |

Table S1: *P*-values for Shapiro-Wilk tests to test the Normality of the NLP measures, for the TAT speech excerpts.

### 3 Relationships between NLP measures and the TLI, PANSS scores and cognitive measures

Table S2 shows the associations between the NLP measures and the TLI (TLI total, TLI positive and TLI negative), PANSS symptoms (PANSS positive, PANSS negative and PANSS general), WRAT IQ and number of years in education. We note that all subjects had data available for the TLI, IQ and number of years in education, whilst 15 CHR-P subjects, 8 FEP patients and no control subjects had PANSS data available.

| NLP Measure | TLI total | TLI positive | TLI negative | PANSS positive | PANSS negative | PANSS general | WRAT IQ | Years education |
|---|---|---|---|---|---|---|---|---|
| No. words | 0.13 (0.96) | 1.6 (0.46) | **-4.7 (0.0010)** | 1.1 (0.63) | -0.38 (0.88) | 0.22 (0.94) | -0.48 (0.86) | -1.2 (0.51) |
| No. sent.s | 0.22 (0.94) | 0.42 (0.88) | -0.54 (0.85) | 1.2 (0.54) | -0.26 (0.94) | 0.54 (0.85) | -2.9 (0.077) | -1.4 (0.47) |
| Mean sent. | 0.0039 (1.00) | 0.89 (0.72) | -2.5 (0.13) | -0.15 (0.96) | -0.59 (0.85) | -0.54 (0.85) | 1.5 (0.46) | -0.13 (0.96) |
| Coherence | -0.48 (0.86) | 0.29 (0.94) | -1.9 (0.39) | -0.37 (0.88) | -1.0 (0.65) | -0.60 (0.85) | 1.2 (0.51) | 0.82 (0.77) |
| Tangentiality | -0.44 (0.88) | 0.26 (0.94) | -1.9 (0.39) | -0.53 (0.85) | 0.0064 (1.00) | -1.2 (0.53) | -0.51 (0.86) | 2.5 (0.13) |
| On-topic | -2.8 (0.077) | -2.0 (0.33) | -1.8 (0.41) | -1.5 (0.46) | -1.8 (0.43) | -0.78 (0.78) | 1.7 (0.46) | 1.5 (0.46) |
| Max. similarity | 1.0 (0.65) | 0.98 (0.65) | -0.027 (1.00) | -1.3 (0.51) | -1.4 (0.49) | -1.3 (0.50) | -1.4 (0.46) | -1.4 (0.47) |
| Ambig. pronouns | 1.1 (0.59) | 1.6 (0.46) | -1.5 (0.46) | 1.5 (0.46) | -0.43 (0.88) | 1.6 (0.46) | -0.12 (0.96) | 0.0083 (1.00) |
| LCC | -0.72 (0.80) | 0.70 (0.81) | **-4.1 (0.0038)** | 0.76 (0.78) | -0.56 (0.85) | 0.083 (0.99) | 1.7 (0.43) | 1.9 (0.39) |
| LSC | -2.1 (0.27) | -0.39 (0.88) | **-5.4 (0.00018)** | -0.21 (0.94) | -1.2 (0.53) | -0.41 (0.88) | 2.6 (0.13) | 1.6 (0.46) |
| LCCr | -0.76 (0.78) | 0.76 (0.78) | **-4.4 (0.0023)** | 0.21 (0.94) | -0.62 (0.85) | -0.56 (0.85) | 1.5 (0.46) | 1.6 (0.46) |
| LSCr | -1.3 (0.51) | 0.042 (1.00) | **-3.6 (0.014)** | 0.59 (0.85) | -0.86 (0.76) | 0.26 (0.94) | 2.9 (0.077) | 2.4 (0.15) |

Table S2: Supplementary Table S1 shows the associations between the NLP measures and the TLI (TLI total, TLI positive and TLI negative), PANSS symptoms (PANSS positive, PANSS negative and PANSS general), WRAT IQ, and number of years in education. Results are shown as $T$-statistics, with FDR corrected $P$-values in brackets (corrected for $12 \times 8 = 96$ multiple comparisons.

### 4 Medication

To assess whether our results were likely to be driven by group differences in medication, we re-calculated the group differences in NLP measures for the TAT pictures excluding the 4 CHR-P subjects and 6 FEP patients who had been prescribed medication. The results are given in Table S3 and are qualitatively similar to the results in the main text. We therefore conclude that group differences in medication did not explain the results observed.

| NLP measure | FEP/CON | CHR-P/CON | FEP/CHR-P |
|---|---|---|---|
| No. words | -1.7 (0.081) | -1.5 (0.12) | -0.64 (0.52) |
| No. sent.s | **2.0 (0.044)** | 0.39 (0.70) | 1.7 (0.082) |
| Mean sent. length | **-3.3 (<0.001)** | -1.5 (0.14) | **-2.2 (0.029)** |
| Coherence | **-3.1 (0.0022)** | **-2.1 (0.034)** | -1.5 (0.14) |
| Tangentiality | -1.1 (0.28) | -0.72 (0.47) | -0.022 (0.98) |
| On-topic | **-3.2 (0.0014)** | **-2.7 (0.0061)** | -1.1 (0.26) |
| Max. similarity | 1.4 (0.16) | 0.60 (0.55) | 1.3 (0.21) |
| Ambig. Pronouns | 0.24 (0.81) | 1.5 (0.15) | -1.2 (0.25) |
| LCC | **-2.9 (0.0033)** | -1.5 (0.13) | **-2.5 (0.011)** |
| LSC | -1.5 (0.14) | -1.5 (0.14) | -0.81 (0.42) |
| LCCr | **-3.1 (0.0022)** | -1.8 (0.077) | **-2.3 (0.021)** |
| LSCr | **-3.1 (0.0017)** | -1.1 (0.27) | **-2.3 (0.021)** |

Table S3: Group differences in NLP measures for the TAT pictures after excluding the 4 CHR-P subjects and 6 FEP patients who had been prescribed medication. The Z-values from Mann-Whitney U-tests are given, with the corresponding $P$-values in brackets.

### 5 Transition to psychosis

We did not observe significant differences in any of the twelve NLP measures between the 8 CHR-P subjects who subsequently transitioned to psychosis and the 16 CHR-P subjects who did not transition; see Table S4.

| NLP measure | $Z$-value | $P$-value |
|---|---|---|
| No. words | -0.031 | 0.98 |
| No. sent.s | -0.49 | 0.62 |
| Mean sent. length | 0.34 | 0.74 |
| Coherence | -0.092 | 0.93 |
| Tangentiality | 1.3 | 0.19 |
| On-topic | -0.46 | 0.65 |
| Max. similarity | 1.4 | 0.17 |
| Ambig. Pronouns | -0.16 | 0.88 |
| LCC | -1.1 | 0.26 |
| LSC | -1.1 | 0.28 |
| LCCr | -0.89 | 0.37 |
| LSCr | -1.4 | 0.15 |

Table S4: Group differences in NLP measures for the CHR-P subjects who did and did not transition to psychosis (Mann-Whitney U-test, for the TAT picture results).

### 6 Inaudible pieces of speech

Inaudible pieces of speech were marked as [?] in the transcripts. We counted the number of inaudible pieces of speech per excerpt, then divided by the total number of words in the excerpt to get number of inaudible pieces of speech per word.

The differences in the number of inaudible pieces of speech per word between groups for the TAT, DCT and free speech recordings are shown in Figure S1 and Table S5. For the TAT there was a significant difference in the number of inaudible pieces of speech per word between the CHR-P and control groups, whilst for the DCT there was a significant difference between the FEP and control groups.

| | TAT | DCT | Free |
|---|---|---|---|
| FEP/CON | 1.3 (P=0.20) | **2.0 (P=0.047)** | 0.41 (P=0.68) |
| FEP/CHR-P | -0.95 (P=0.34) | 1.7 (P=0.086) | -0.24 (P=0.81) |
| CHR-P/CON | **2.5 (P=0.014)** | 1.1 (P=0.26) | 0.36 (P=0.72) |

Table S5: Differences in the number of inaudible pieces of speech per word between the TAT, DCT and free speech recordings, calculated using the Mann-Whitney U-test.

We therefore tested whether the previously identified group differences in the NLP metrics from the TAT and DCT remained significant when controlling for the number of inaudible pieces of speech per word. To that end we used a Generalized Additive Model for Location, Scale and Shape (GAMLSS) with a gamma distribution [2], including number of inaudible pieces of speech per word as a co-variate. Results are shown in Tables S6 and S7. All previously identified significant group differences remained significant, apart from the group differences in total number of words between FEP patients and control subjects, in both the TAT and DCT speech excerpts. We also note that we were unable to test for a group difference in ambiguous pronoun count between the FEP patients and control subjects in the DCT excerpts using the GAMLSS model due to some subjects having ambiguous pronoun counts of 0 (which is incompatible with using a gamma distribution).

| | FEP/CON | CHR-P/CON | FEP/CHR-P |
|---|---|---|---|
| No. words | -1.8 (P=0.084) | N/A | N/A |
| No. sentences | 2.4 (P=0.024) | N/A | N/A |
| Sentence length | -3.6 (P=0.0014) | N/A | N/A |
| Coherence | -4.5 (P< 0.001) | -2.3 (P=0.029) | N/A |
| Tangentiality | N/A | N/A | N/A |
| On-topic | -5.0 (P< 0.001) | -3.4 (P=0.0018) | N/A |
| Max. similarity | N/A | N/A | N/A |
| Ambig. pronouns | N/A | N/A | N/A |
| LCC | -3.9 (P< 0.001) | N/A | -3.6 (P< 0.001) |
| LSC | N/A | N/A | N/A |
| LCCr | -3.8 (P< 0.001) | N/A | -3.3 (P=0.0024) |
| LSCr | -3.4 (P=0.0025) | N/A | -2.8 (P=0.0091) |

Table S6: Group differences in NLP metrics from the TAT, assessed using a GAMLSS model with a gamma distribution, controlling for number of inaudible pieces of speech per word as a co-variate.

The differences in the number of inaudible pieces of speech per word between the TAT, DCT and free speech recordings are shown in Figure S2 and Table S8.

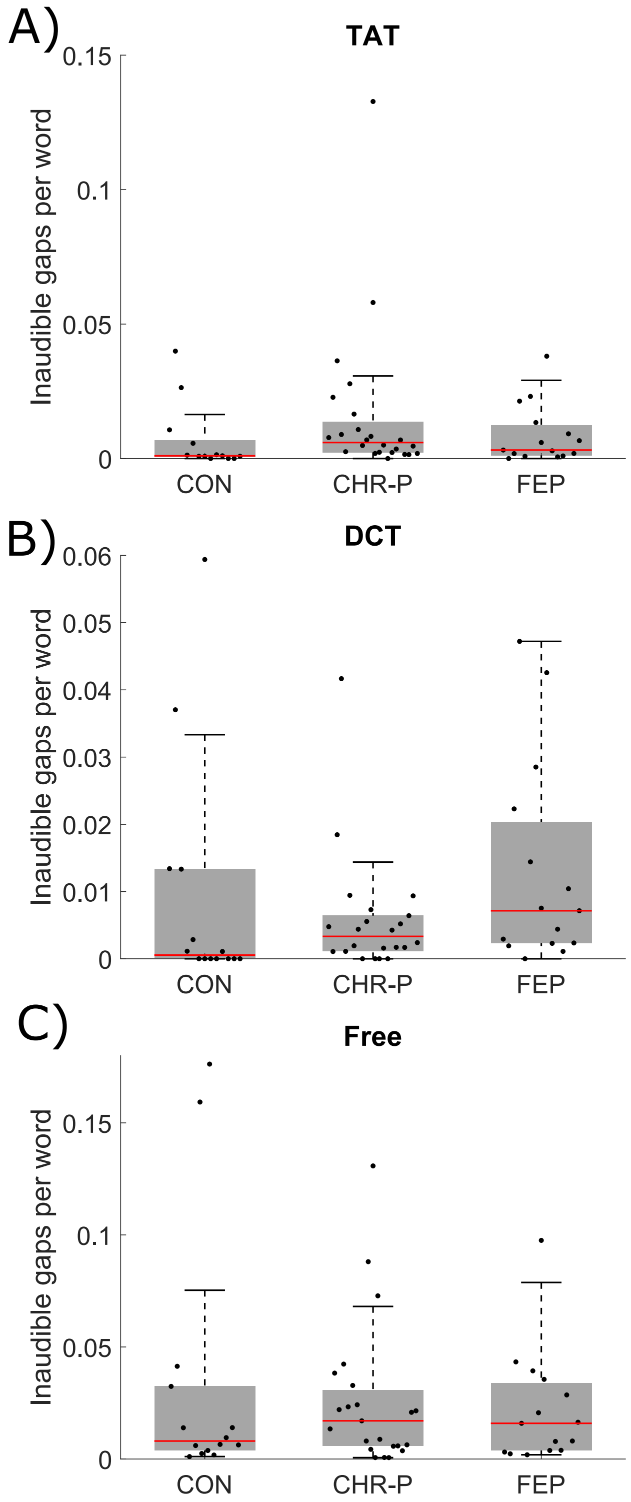


Figure S1: Group differences in number of inaudible pieces of speech per word, for the A) TAT, B) DCT and C) free speech excerpts.

| | FEP/CON | CHR-P/CON | FEP/CHR-P |
|---|---|---|---|
| No. words | -0.28 (P=0.78) | N/A | N/A |
| No. sentences | N/A | N/A | N/A |
| Sentence length | -2.4 (P=0.025) | N/A | -2.2 (P=0.035) |
| Coherence | -3.6 (P=0.0015) | N/A | -2.3 (P=0.028) |
| Tangentiality | N/A | N/A | N/A |
| On-topic | -4.0 (P< 0.001) | N/A | -2.2 (P=0.034) |
| Max. similarity | N/A | N/A | N/A |
| Ambig. pronouns | * | N/A | N/A |
| LCC | -3.5 (P=0.0020) | N/A | -2.5 (P=0.019) |
| LSC | -3.3 (P=0.0026) | N/A | -2.1 (P=0.043) |
| LCCr | -3.0 (P=0.0062) | N/A | N/A |
| LSCr | -4.3 (P< 0.001) | -2.6 (P=0.026) | -3.0 (P=0.0055) |

Table S7: Group differences in NLP metrics from the DCT, assessed using a GAMLSS model with a gamma distribution, controlling for number of inaudible pieces of speech per word as a co-variate. *We note that we were unable to test for a group difference in ambiguous pronoun count between the FEP patients and control subjects- see text for details.

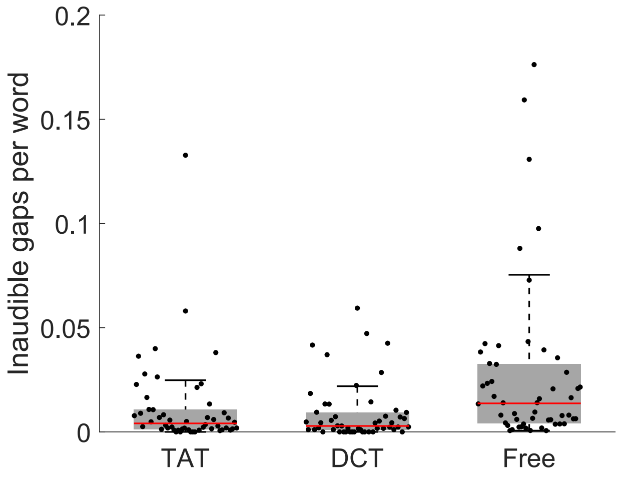


Figure S2: Number of inaudible pieces of speech per word, for the TAT, DCT and free speech excerpts.

| | Z-value | P-value |
|---|---|---|
| TAT/DCT | 0.81 | 0.42 |
| TAT/Free | -3.4 | < 0.001 |
| DCT/Free | -4.0 | < 0.001 |

Table S8: Differences in the number of inaudible pieces of speech per word between the TAT, DCT and free speech recordings, calculated using the Mann-Whitney U-test.

### References

[1] P. Shrestha, "Detailed Procedure of Thematic Apperception test," 2017.

[2] R. Rigby, D. Stasinopoulos, and P. Lane, "Generalized additive models for location, scale and shape," *J. R. Stat. Soc. Ser. C Appl. Stat.*, vol. 54, pp. 507–554, 2005.
